# Timing matters: when do primiparous and multiparous pregnant women initiate folic acid supplementation, and who initiates it too late?

**DOI:** 10.64898/2026.09.25.26363991

**Authors:** Utibe S. Ebong, Joanne E. Given, Frank A. Casey, Maria Loane, Helen Dolk

**Affiliations:** School of Medicine, Faculty of Life and Health Sciences, Ulster University.; Institute of Nursing and Health Research, Faculty of Life and Health Sciences, Ulster University.

**Author notes:** **Corresponding author:** Professor Helen Dolk, School of Medicine, Faculty of Life and Health Sciences, Ulster University, 2-24 York Street, Belfast BT15 1ED.

**Keywords:** Folic Acid, Parity, Pregnancy, Antenatal Care, Preconception Care

## Abstract

**Background:** Women are recommended to take folic acid supplements (FAS) periconceptionally to prevent neural tube defects, but the low levels of FAS uptake are poorly understood. This study aimed to determine how parity, pregnancy planning and other maternal factors affect the timing of FAS initiation.

**Method:** We used data from a representative sample of 966 pregnant women from the Northern Ireland Baby Hearts study. Questionnaire responses on health behaviours, including FAS, were linked to maternity and area-based deprivation data. Logistic regression assessed determinants of FAS initiation preconception. Time-varying Cox regression assessed associations between initiation postconception and pregnancy-related milestones.

**Results:** Forty-one per cent of women initiated FAS preconception. Lower odds of preconception initiation were associated with multiparity (adjusted OR 0.47 (0.32 – 0.68)), unplanned pregnancy (adjusted OR 0.08 (0.05 – 0.13)), deprivation and consuming a low folate diet. Higher odds were associated with fertility treatment and previous pregnancy loss. Forty-seven per cent of women initiated FAS postconception, mainly after pregnancy recognition (median 5 weeks), including 34.1% of those who planned their pregnancy. For primiparous women with unplanned pregnancies, the first health service contact (median 7 weeks) further increased FAS initiation.

**Conclusion:** Many women, even multiparous women with previous pregnancy experience and women who planned their pregnancies, are initiating FAS after pregnancy recognition. This shows awareness of FAS benefits, but is too late to prevent neural tube defects. Public health interventions need to go beyond awareness to address motivation and opportunity for FAS, and pay more attention to multiparous women.

## INTRODUCTION

Although neural tube defects (NTDs) are mainly preventable, it is estimated that each year, 260,100 pregnancies worldwide are affected.^1^ Cases of NTDs either result in fatal outcomes when severe^2^ or life-long disability for surviving infants.^3^ The neural tube closes within 28 days after conception and requires sufficient folate levels.^4,5^ Across Europe, women are advised to take 400 micrograms of folic acid starting at least one month before pregnancy or when planning a pregnancy.^6^ Preconception folic acid supplementation (FAS) varies widely across Europe and often falls below recommendations.^7^ As a complementary strategy, staple foods may be fortified with folic acid. While many countries outside Europe mandate folic acid food fortification, fortification is largely voluntary across Europe.^8^ The impact of folic acid food fortification in reducing NTDs is well documented.^9^ The UK government has mandated the fortification of flour with folic acid from December 13, 2026.

The unplanned nature of many pregnancies has been identified as a major barrier to FAS initiation preconception.^12,13^ Actively planning to become pregnant often prompts women to adopt healthy behaviours, including FAS.^12,14^ During preconception care, early initiation of FAS is prioritised as an important counselling topic,^15^ leading to higher rates of preconception FAS initiation.^16,17^ Given their likely exposure to information on FAS from previous pregnancies, multiparous women might be expected to have higher FAS uptake preconception. However, in a systematic review and meta-analysis, we found that multiparous women globally were less likely than primiparous women to initiate FAS preconception, even after accounting for pregnancy planning and other factors.^18^ Many women who do not initiate FAS preconception may initiate FAS postconception; however, we know little of the timing of this initiation postconception and its determinants. This study aimed to determine how parity, pregnancy planning and other maternal factors affect the timing of FAS initiation.

## METHODS

### Study design and participants

This study was based on an analysis of the control group of the Northern Ireland (NI) Baby Hearts case-control study, ^19^ which aimed to investigate the potential environmental causes of congenital heart disease. Controls were 966 resident mothers of babies without congenital heart disease, recruited from 2014 to 2016 during clinical screening (anomaly scan) appointments at 18–20 gestational weeks. The control sample was representative of pregnant women in NI during the study period.^19^

The NI Baby Hearts study linked prospectively collected questionnaire data to maternity records from the NI Maternity System (NIMATS) and area-based deprivation data from the 2010 NI Multiple Deprivation Measure. Area-based deprivation was based on women’s postcode of residence as documented on their NIMATS records at the first antenatal appointment. The methodology used has been previously explained.^19^ Information about the questionnaire is publicly available on <u>The Baby Hearts Study (Northern Ireland) Database —Ulster University</u> and extracts are given in Table S1.

### Main Outcome Measures

Folic acid uptake was assessed via the questionnaire, where women were asked questions about FAS (*“Did you take folic acid at any time during the THREE MONTHS BEFORE you became pregnant and/or during the FIRST THREE MONTHS of your pregnancy?”* with the following response options: (i) Yes, on its own (ii) Yes, in multivitamins or as part of an iron supplement (iii) No (iv) I don’t know. Timing of FAS initiation (whether on its own or as part of a multivitamin) was obtained from the question *“How many weeks pregnant were you when you started to take folic acid?”*

### Definition of parity, pregnancy planning and pregnancy-related milestones

We obtained parity data from NIMATS, which defined parity as the number of previous live births or stillbirths at 24 gestational weeks or more. Primiparous women were thus defined as women having their first viable pregnancy (≥24 weeks), whereas multiparous women were defined as pregnant women with a previously viable pregnancy regardless of pregnancy outcome.

Pregnancy planning was defined based on responses to: *“How long were you trying to get pregnant?”* Women who reported they were not trying to become pregnant were categorised as having an unplanned pregnancy.

Pregnancy-related milestones included the timing of pregnancy recognition, first contact with a healthcare professional, and the antenatal booking appointment. We defined pregnancy recognition as the period from the gestational week when a woman first suspected she might be pregnant to the week when the pregnancy was confirmed. Suspecting pregnancy was drawn from questionnaire responses to the question: *“How many weeks pregnant were you when you first thought you might be pregnant?”* Confirming pregnancy was based on questionnaire responses to the following: *“How many weeks pregnant were you when you had your pregnancy confirmed?”*

Contact with a healthcare professional was drawn from questionnaire responses to the question: *“How many weeks were you when you first saw a health professional about your pregnancy?”* We obtained the gestational week at antenatal booking from NIMATS.

### Covariates

Women were asked about consumption of folate-rich foods in the three months before pregnancy, such as leafy vegetables, broccoli, chickpeas, and liver. The frequency of consumption of these and other foods in the three months before pregnancy was used to allocate women to one of three dietary classes in latent class analysis:^20^ Type 1: moderate fruits and vegetables, Type 2: varied diet with high fruits and vegetables, and Type 3: poor diets with low fruits and vegetables.

Selectively buying folic acid-fortified foods was assessed from questionnaire responses to the question: *“Do you buy cereals, breads, cereal bars or spreads fortified with folic acid?”* with the following response options: (i) Yes, since before I became pregnant (ii) Yes, since I became pregnant (iii) No (iv) Don’t know. A second question asked about eating fortified breads, breakfast cereals, cereal bars and spreads during the first three months of pregnancy. The questionnaire also included a list of fortified foods sold in NI at the time, so women could identify which they consumed (whether or not they knew these foods were fortified). They were then categorised based on the number of fortified foods consumed during the first three months of pregnancy: none, one food type, or two or more types.

Higher risk of NTDs referred to women with pregestational diabetes, epilepsy, or obesity (as per NI guidelines).^21^ Of the women in this category, 96% were women with obesity and 4% were women with epilepsy and diabetes. Variables used in this study are defined in Table S1.

### Statistical analyses

We used the chi-square test to assess variations in categorical variables, the t-test for differences in continuous variables, and the Mann-Whitney test for differences in median values. All tests excluded women with missing values on the variable tested; the number of women with missing values for each variable is shown in the relevant tables.

Logistic regression analysis predicted determinants of FAS initiation preconception. A directed acyclic graph (DAG) was used to visualise causal pathways and identify potential confounding variables for adjustment in the regression model.^22^ Findings from previous studies were used to identify relevant variables for the DAG and characterise their relationships (Figure S1). The DAG identified eight potential confounding factors: (i) Age, (ii) Deprivation, (iii) Education, (iv) Marital status, (v) Occupation, (vi) Previous loss/Abortion, (vii) Pregnancy planning and (viii) Race/Ethnicity/Nationality. These were used to adjust the logistic regression model. A stratified analysis using parity and pregnancy planning was used to assess the interaction between parity, pregnancy planning and the determinants of preconception FAS initiation. Unadjusted and adjusted odds ratios (ORs) are reported with 95% confidence intervals (CI).

Kaplan-Meier survival analysis was used to plot the timing of FAS initiation. The week when FAS was initiated was coded as the event, and gestational week 14 as the censoring time. Kaplan-Meier cumulative survival curves were generated by parity and pregnancy planning status, using median gestational ages for pregnancy-related milestones as vertical reference lines. A time-varying Cox regression model was used to examine the effect of pregnancy-related milestones on postconception FAS initiation on an individual basis, by parity and pregnancy planning (limited to women who initiated FAS postconception). Hazard ratios (HR) and 95% CI for the associations are reported. Analyses were conducted using Stata version 18.^23^

### Ethics approval

This study was approved by Ulster University’s Biomedical Sciences Ethics Filter Committee (FCBMS-24-016). The NI Baby Hearts study was approved by the Office for Research Ethics Committees Northern Ireland (14NI0027).

## RESULTS

### Description of study participants by parity

Of the 966 women in the study, 393 (40.7%) were primiparous, and 573 (59.3%) were multiparous. Two-thirds (66.9%) of the women planned their pregnancy, with no difference by parity (Table 1). Multiparous women were more likely than primiparous women to be older, married, with basic education, and not employed (Table 1). Multiparous women were more likely to have a history of pregnancy loss, less likely to have had infertility treatment, and more likely to be at higher risk of having a child with NTD because of the higher prevalence of obesity (BMI>30) (Table 1).

**Table 1:** Sociodemographic and other risk factors by parity. Alt text: Descriptive table showing the sociodemographic characteristics and other risk factors, disaggregated by parity, among pregnant women in Northern Ireland.

|  | **Primiparous women**<br>**(n = 393)** | **Multiparous women**<br>**(n = 573)** | **Total**<br>**(n = 966)** | **p-value** |
| --- | --- | --- | --- | --- |
| Maternal age |  |  |  |  |
| <25 | 106 (26.9%) | 52 (9.1%) | 158 (16.4%) | < 0.001 |
| 25-29 | 117 (29.8%) | 139 (24.3%) | 256 (26.5%) |  |
| 30-34 | 128 (32.6%) | 222 (38.7%) | 350 (36.2%) |  |
| 35+ | 42 (10.7%) | 160 (27.9%) | 202 (20.9%) |  |
| Marital status |  |  |  |  |
| Married/Civil partnership | 207 (52.7%) | 377 (65.8%) | 584 (60.5%) | < 0.001 |
| In a steady relationship/Living with someone | 156 (39.7%) | 155 (27.1%) | 311 (32.2%) |  |
| Single/Separated/Divorced/Widowed | 30 (7.6%) | 41 (7.1%) | 71 (7.4%) |  |
| Maternal education |  |  |  |  |
| Left with no or basic qualifications | 62 (15.8%) | 131 (22.9%) | 193 (20.0%) | 0.03 |
| Higher secondary / Technical college | 150 (38.2%) | 201 (35.1%) | 351 (36.3%) |  |
| University degree | 181 (46.0%) | 240 (41.9%) | 421 (43.6%) |  |
| Missing | - | 1 (0.2%) | 1 (0.1%) |  |
| Maternal occupation |  |  |  |  |
| Unemployed | 22 (5.6%) | 46 (8.0%) | 68 (7.0%) | < 0.001 |
| Employed | 352 (89.6%) | 417 (72.8%) | 769 (79.6%) |  |
| Homemakers | 4 (1.0%) | 104 (18.2%) | 108 (11.2%) |  |
| Students | 15 (3.8%) | 5 (0.9%) | 20 (2.1%) |  |
| Missing | - | 1 (0.2%) | 1 (0.1%) |  |
| Area-based socioeconomic deprivation |  |  |  |  |
| 1 (most deprived) | 77 (19.6%) | 119 (20.8%) | 196 (20.3%) | 0.89 |
| 2 | 78 (19.9%) | 122 (21.3%) | 200 (20.7%) |  |
| 3 | 84 (21.4%) | 128 (22.3%) | 212 (21.9%) |  |
| 4 | 80 (20.4%) | 111 (19.4%) | 191 (19.8%) |  |
| 5 (least deprived) | 64 (16.3%) | 83 (14.5%) | 147 (15.2%) |  |
| Missing | 10 (2.5%) | 10 (1.8%) | 20 (2.1%) |  |
| Pregnancy planning |  |  |  |  |
| Did not plan to become pregnant | 136 (34.6%) | 184 (32.1%) | 320 (33.1%) | 0.42 |
| Planned to get pregnant | 257 (65.4%) | 389 (67.9%) | 646 (66.9%) |  |
| Infertility treatment |  |  |  |  |
| None | 316 (80.4%) | 526 (91.8%) | 842 (87.1%) | < 0.001 |
| Investigations only | 18 (4.6%) | 11 (1.9%) | 29 (3.0%) |  |
| Treated for infertility | 35 (8.9%) | 12 (2.1%) | 47 (4.9%) |  |
| Missing | 24 (6.1%) | 24 (4.2%) | 48 (5.0%) |  |
| Previous pregnancy loss |  |  |  |  |
| None | 310 (78.8%) | 317 (55.3%) | 627 (64.9%) | < 0.001 |
| 1 | 59 (15.0%) | 165 (28.8%) | 224 (23.2%) |  |
| 2 or more | 12 (3.1%) | 75 (13.1%) | 87 (9.0%) |  |
| Missing | 12 (3.1%) | 16 (2.8%) | 28 (2.9%) |  |

|  | Primiparous women<br>(n = 393) | Multiparous women<br>(n = 573) | Total<br>(n = 966) | p-value |
| --- | --- | --- | --- | --- |
| Chronic conditions□ |  |  |  |  |
| No | 240 (61.1%) | 347 (60.6%) | 587 (60.8%) | 0.87 |
| Yes | 153 (38.9%) | 226 (39.4%) | 379 (39.2%) |  |
| Body mass index at booking |  |  |  |  |
| Underweight | 2 (0.5%) | 9 (1.6%) | 11 (1.1%) | 0.04 |
| Healthy weight | 201 (51.2%) | 257 (44.9%) | 458 (47.4%) |  |
| Overweight | 112 (28.5%) | 157 (27.4%) | 269 (27.9%) |  |
| Obese | 68 (17.3%) | 134 (23.4%) | 202 (20.9%) |  |
| Missing | 10 (2.5%) | 16 (2.8%) | 26 (2.7%) |  |
| Pregnant women with a higher risk of NTD<sup>§</sup> |  |  |  |  |
| No | 319 (81.2%) | 433 (75.6%) | 752 (77.8%) | 0.04 |
| Yes | 74 (18.8%) | 140 (24.4%) | 214 (22.2%) |  |

<sup>§</sup> Higher risk of NTDs referred to women with pregestational diabetes, epilepsy, or obesity (as per NI guidelines).<sup>21</sup> This category was mostly made up of women with obesity (96%). The remaining 4% were women with epilepsy and diabetes.
□ Chronic conditions were diabetes, asthma, anxiety/stress, depression, bipolar disorder, obsessive-compulsive disorder, panic disorder, other mental health problems, raised blood pressure, epilepsy, obesity, anorexia, anaemia, clotting disorder, and heart disease acquired in adulthood.

The median gestational ages (GA) at which women suspected they were pregnant and when they confirmed they were pregnant were week 4 (IQR 3-5) and week 5 (IQR 4-6), respectively, which did not differ by parity. Pregnancy recognition was therefore from week 4 to 5. The median GA at which women first contacted a health professional about the pregnancy was week 7 (IQR 5-8). Primiparous women spoke to a health professional earlier than multiparous women (weeks 6 and 7, respectively; p <0.001). The median GA at which women had their antenatal booking appointment was 11 weeks (IQR 10-12), and this did not differ by parity.

### Preconceptional initiation of folic acid supplementation

Preconceptional FAS initiation was reported by 40.8% of the women. After adjusting for confounders, the odds of initiating FAS preconception were 53% lower among multiparous than primiparous women (aOR 0.47, 95% CI 0.32 – 0.68; Table 2). The odds of initiation preconception were 92% lower among women with an unplanned pregnancy compared to those with a planned pregnancy (aOR 0.08, 95% CI 0.05 – 0.13; Table 2). Even among those with planned pregnancies, 34.1% initiated FAS postconception and 8.2% did not take FAS at all (Table S3). The observed effect of pregnancy planning on FAS initiation preconception did not differ by parity (Table S4).

**Table 2:** Determinants of preconception folic acid initiation with unadjusted and adjusted effect estimates. Alt text: Descriptive table showing the sociodemographic determinants of preconceptional initiation of folic acid supplementation among pregnant women in Northern Ireland.

| Sociodemographic characteristics | Preconception<br>(n = 394) | Postconception<br>or No uptake<br>(n = 572) | Unadjusted OR<br>(95% CI) | Adjusted OR<sup>a</sup><br>(95% CI) |
| --- | --- | --- | --- | --- |
| Parity |  |  |  |  |
| Primiparous women | 176 (44.8%) | 217 (55.2%) | Reference | Reference |
| Multiparous women | 218 (38.1%) | 355 (61.9%) | 0.76 (0.58 – 0.98) | 0.47 (0.32 – 0.68) |
| Pregnancy planning |  |  |  |  |
| Did not plan | 21 (6.6%) | 299 (93.4%) | 0.05 (0.03 – 0.08) | 0.08 (0.05 – 0.13) |
| Planned to get pregnant | 373 (57.7%) | 273 (42.3%) | Reference | Reference |
| Maternal age |  |  |  |  |
| ≤24 | 20 (12.7%) | 138 (87.3%) | 0.14 (0.08 – 0.23) | 0.67 (0.35 – 1.28) |
| 25-29 | 88 (34.4%) | 168 (65.6%) | 0.49 (0.36 – 0.69) | 0.76 (0.51 – 1.14) |
| 30-34 | 180 (51.4%) | 170 (48.6%) | Reference | Reference |
| 35+ | 106 (52.5%) | 96 (47.5%) | 1.04 (0.74 – 1.48) | 1.28 (0.83 – 1.96) |
| Marital status |  |  |  |  |
| Married / Civil partnership | 315 (53.9%) | 269 (46.1%) | Reference | Reference |
| Steady relationship / Living with someone | 72 (23.1%) | 239 (76.9%) | 0.26 (0.19 – 0.35) | 0.67 (0.44 – 1.01) |
| Single/Separated/Divorced/Widowed | 7 (9.9%) | 64 (90.1%) | 0.09 (0.04 – 0.21) | 0.39 (0.15 – 1.04) |
| Maternal education |  |  |  |  |
| Left with no or basic qualifications | 53 (27.5%) | 140 (72.5%) | 0.30 (0.21 – 0.44) | 0.94 (0.57 – 1.55) |
| Higher secondary / Technical college | 107 (30.5%) | 244 (69.5%) | 0.35 (0.26 – 0.47) | 0.72 (0.49 – 1.05) |
| University degree | 234 (55.6%) | 187 (44.4%) | Reference | Reference |
| Missing | -- | 1 (100.0%) |  |  |
| Maternal occupation |  |  |  |  |
| Unemployed | 6 (8.8%) | 62 (91.2%) | 0.11 (0.05 – 0.26) | 0.31 (0.17 – 0.83) |
| Employed | 357 (46.4%) | 412 (53.6%) | Ref | Ref |
| Homemakers | 31 (28.7%) | 77 (71.3%) | 0.47 (0.29 – 0.72) | 0.54 (0.31 – 0.92) |
| Students | -- | 20 (100.0%) | -- | -- |
| Missing | -- | 1 (100.0%) |  |  |
| Area-based deprivation |  |  |  |  |
| 1 (most deprived) | 54 (27.6%) | 142 (72.4%) | 0.32 (0.20 – 0.50) | 0.47 (0.26 – 0.83) |
| 2 | 75 (37.5%) | 125 (62.5%) | 0.50 (0.33 – 0.78) | 0.54 (0.32 – 0.93) |
| 3 | 102 (48.1%) | 110 (51.9%) | 0.78 (0.51 – 1.18) | 0.82 (0.48 – 1.39) |
| 4 | 75 (39.3%) | 116 (60.7%) | 0.54 (0.35 – 0.84) | 0.52 (0.31 – 0.89) |
| 5 (least deprived) | 80 (54.4%) | 67 (45.6%) | Reference | Reference |
| Missing | 8 (40.0%) | 12 (60.0%) |  |  |
| Selectively bought fortified foods |  |  |  |  |
| No | 117 (37.3%) | 197 (62.7%) | Reference | Reference |
| Yes, since before I became pregnant | 148 (44.3%) | 186 (55.7%) | 1.34 (0.98 – 1.84) | 1.20 (0.81 – 1.79) |
| Yes, since I became pregnant | 14 (25.0%) | 42 (75.0%) | 0.56 (0.29 – 1.07) | 0.89 (0.41 – 1.93) |
| I don't know | 115 (43.9%) | 147 (56.1%) | 1.32 (0.94 – 1.84) | 1.42 (0.94 – 2.17) |
| Ate fortified foods during the first trimester |  |  |  |  |
| None | 87 (45.1%) | 106 (54.9%) | Reference | Reference |
| One type | 152 (40.5%) | 223 (59.5%) | 0.83 (0.59 – 1.18) | 0.89 (0.57 – 1.39) |
| Two types or more types | 155 (38.9%) | 243 (61.1%) | 0.78 (0.55 – 1.10) | 0.91 (0.59 – 1.41) |
| Diet in the 3 months before pregnancy |  |  |  |  |
| Type 1 – Moderate diet | 158 (41.8%) | 220 (58.2%) | 0.64 (0.47 – 0.86) | 0.69 (0.47 – 0.98) |
| Type 2 – Varied diet with high fruits & veg | 173 (53.1%) | 153 (46.9%) | Reference | Reference |
| Type 3 – Poor diet with low fruits & veg | 63 (24.2%) | 197 (75.8%) | 0.28 (0.19 – 0.40) | 0.47 (0.29 – 0.73) |
| Missing | -- | 2 (100.0%) |  |  |
| Infertility treatment |  |  |  |  |
| None | 316 (37.5%) | 526 (62.5%) | Reference | Reference |
| Investigations only | 15 (51.7%) | 14 (48.3%) | 1.78 (0.85 – 3.74) | 0.82 (0.36 – 1.87) |
| Treated for infertility | 38 (80.8%) | 9 (19.2%) | 7.03 (3.35 – 14.73) | 4.51 (1.94 – 10.50) |
| Missing | 25 (52.1%) | 23 (47.9%) |  |  |
| Previous pregnancy loss |  |  |  |  |
| None | 235 (37.5%) | 392 (62.5%) | Reference | Reference |
| 1 | 98 (43.7%) | 126 (56.3%) | 1.29 (0.95 – 1.77) | 1.46 (0.98 – 2.15) |
| 2 or more | 45 (51.7%) | 42 (48.3%) | 1.79 (1.14 – 2.80) | 2.05 (1.15 – 3.66) |
| Missing | 16 (57.1%) | 12 (42.9%) |  |  |
| Chronic conditions |  |  |  |  |
| No | 254 (43.3%) | 333 (56.7%) | Reference | Reference |
| Yes | 140 (36.9%) | 239 (63.1%) | 0.77 (0.59 – 1.00) | 0.96 (0.69 – 1.34) |
| Pregnant women with a higher risk of NTDs |  |  |  |  |
| No | 312 (41.5%) | 440 (58.5%) | Reference | Reference |
| Yes | 82 (38.3%) | 132 (61.7%) | 0.88 (0.64 – 1.19) | 1.24 (0.84 – 1.85) |
| Body mass index at booking appointment |  |  |  |  |
| Underweight | 3 (27.3%) | 8 (72.7%) | 0.49 (0.13 – 1.89) | 0.53 (0.09 – 2.82) |
| Healthy weight | 197 (43.0%) | 261 (57.0%) | Reference | Reference |
| Overweight | 103 (38.3%) | 166 (61.7%) | 0.82 (0.60 – 1.12) | 0.78 (0.53 – 1.14) |
| Obese | 75 (37.1%) | 127 (62.9%) | 0.78 (0.56 – 1.09) | 1.07 (0.69 – 1.66) |
| Missing | 16 (61.5%) | 10 (38.5%) |  |  |
| <sup>a</sup> Adjusted for covariates identified in the DAG: (i) Age, (ii) Deprivation, (iii) Education, (iv) Marital status, (v) Occupation, (vi) Pregnancy planning, (vii) Previous pregnancy loss, and (viii) Ethnicity |  |  |  |  |

FAS initiation preconception was reported by 38.3% of women at higher risk of NTDs (Table 2). Women at a higher risk of NTDs had similar odds of initiating FAS preconception to low-risk women (aOR 1.24, 95% CI 0.84 – 1.85; Table 2). Despite planning for the pregnancy, 44.4% initiated FAS postconception or did not take FAS at all (Table S3). Among high-risk women who initiated FAS preconception, 13.4% took the recommended 5mg dose, whereas 86.6% took the normal (400mcg) dose.

Lower odds of preconceptional FAS were associated with living in a more deprived area, younger age, and not being employed (Table 2). Two or more previous pregnancy losses and having received infertility treatment were associated with higher preconceptional FAS (Table 2). The determinants did not differ by parity in their effect on preconceptional FAS initiation except for infertility treatment (Table S4). Among women who received infertility treatment, primiparous women were more likely than multiparous women to initiate FAS preconception (33 out of 35 primiparous women, vs 5 out of 12 multiparous women; Table S4).

### Folic acid (FA) fortified food consumption and folate-rich diets

Women with poor diets (diets low in folate) had significantly lower odds of initiating FAS preconception than women with varied diets (Table 2). About a third (34.6%) ate one or more food types they knew to be FA-fortified in the three months before pregnancy, and a further 5.8% started doing so when they were pregnant. Intentionally eating FA-fortified foods preconception was associated with pregnancy planning, regardless of parity (Table S3). FA-fortified cereals, breads, cereal bars, or spreads, as identified from a list of food brands, were consumed by 80.0% of women during the first trimester. Total fortified food consumption in the first trimester did not vary significantly by parity or pregnancy planning status (Table S3).

### Postconceptional initiation of folic acid supplementation

Close to half (47.2%) of women initiated FAS postconception, and 12.0% were non-users of FAS. Among women with a planned pregnancy, 25.7% of primiparous and 39.6% of multiparous women initiated FAS postconception despite planning for the pregnancy (Table S3). Figure 1 shows cumulative FAS uptake by gestational week of initiation. At week 1, 65.8% of primiparous and 53.9% of multiparous women with planned pregnancies had already initiated FAS. By week 4 (onset of pregnancy recognition), 71.9% of primiparous and 71.2% of multiparous women with planned pregnancies had initiated FAS, levelling the difference by parity. Among women with an unplanned pregnancy, there was a steeper rise in FAS initiation at pregnancy recognition (median weeks 4 to 5). FAS initiation continued to rise for women with an unplanned pregnancy, particularly among primiparous women, after contact with health professionals (median week 7). Antenatal booking did not improve FAS initiation, as uptake was already quite high by this time.

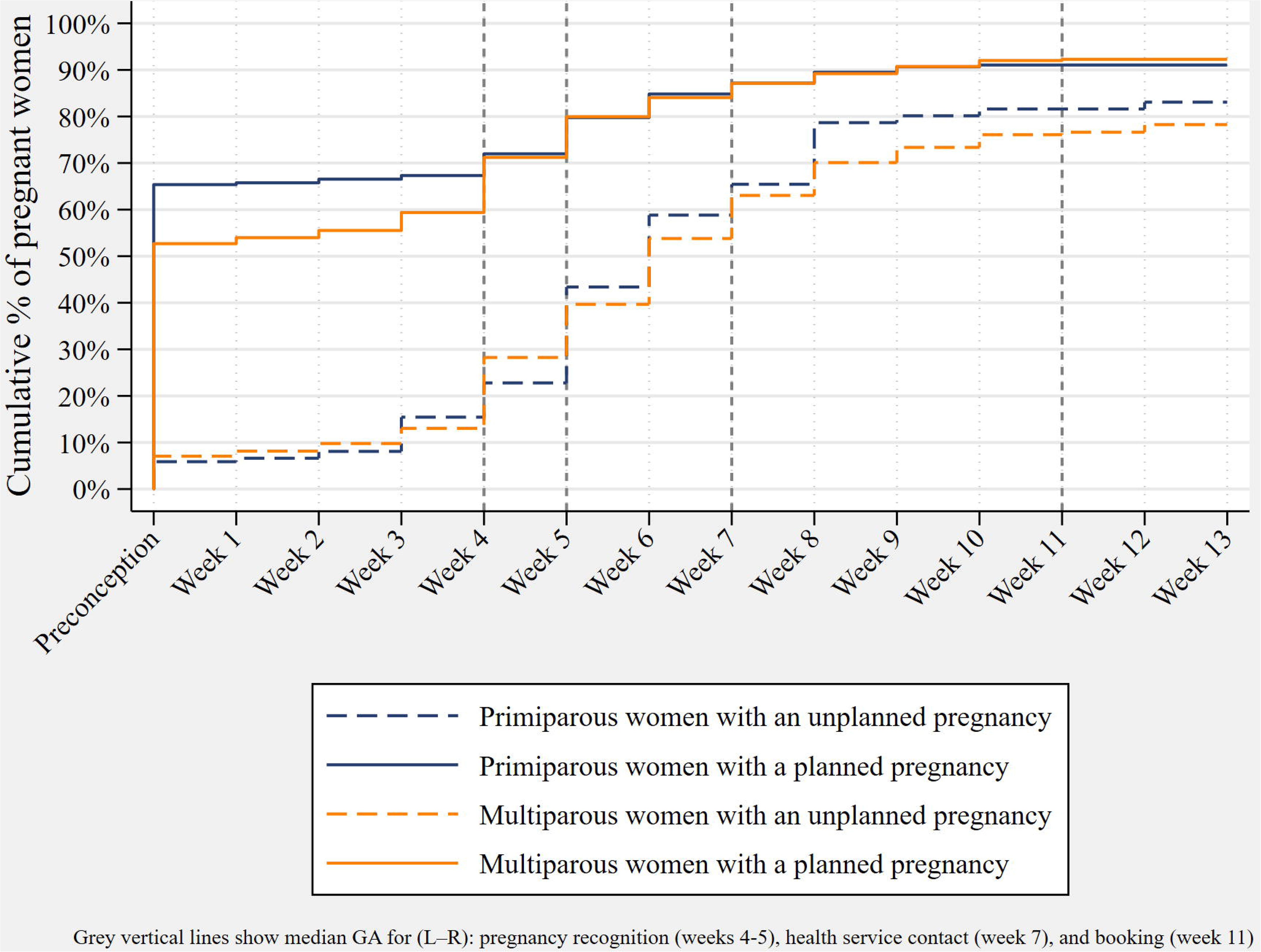

Time-varying Cox regression confirmed that the features found in the Kaplan-Meier plot were statistically significant (Table 3). The greatest increase in FAS initiation postconception occurred after pregnancy recognition, regardless of parity or pregnancy planning status. The first healthcare visit further increased FAS initiation postconception among primiparous women with unplanned pregnancies.

**Table 3:** Hazard ratios showing the association between postconceptional folic acid initiation and pregnancy-related milestones, by parity and pregnancy planning. Alt text: Descriptive table showing the effect of pregnancy-related milestones on postconceptional initiation of folic acid supplementation among pregnant women in Northern Ireland.

| Pregnancy-related milestones | Primiparous women |  | Multiparous women |  | Interaction p-value |
| --- | --- | --- | --- | --- | --- |
|  | Planned pregnancy Unadjusted HR (95 % CI) | Unplanned pregnancy Unadjusted HR (95 % CI) | Planned pregnancy Unadjusted HR (95 % CI) | Unplanned pregnancy Unadjusted HR (95 % CI) |  |
| Before recognition<br>After recognition | Reference<br>18.19 (6.38 – 51.83) | Reference<br>28.45 (8.76 – 92.32) | Reference<br>17.19 (8.48 – 34.81) | Reference<br>25.87 (11.09 – 60.33) | 0.81 |
| Before contact with HCP<br>After contact with HCP | Reference<br>1.57 (0.91 – 2.69) | Reference<br>3.21 (2.17 – 4.76) | Reference<br>1.43 (0.97 – 2.09) | Reference<br>1.57 (1.05 – 2.34) | 0.01 |
| Before booking<br>After booking | Reference<br>0.41 (0.05 – 3.36) | Reference<br>0.62 (0.21 – 1.83) | Reference<br>0.85 (0.31 – 2.36) | Reference<br>0.58 (0.17 – 1.92) | 0.90 |
| *Recognition: Pregnancy recognition*<br>*HCP: Healthcare professional*<br>*Booking: Antenatal booking appointment* |  |  |  |  |  |

### Sources of FAS

Overall, folic acid users in this study bought FAS over the counter (63.8%). Other sources were prescriptions (34.0%) and the Healthy Start scheme (2.2%) – a UK government programme providing support to means-tested pregnant women and young families. More primiparous than multiparous women received FAS prescriptions and free multivitamins from Healthy Start (37.6% vs 31.5% and 3.5% vs 1.4%, respectively; p=0.03).

FAS sources also differed between preconception and postconception users. More preconception users (75.6%) than postconception users (53.5%) bought FAS over the counter. However, more postconception users (43.4%) than preconception users (23.1%) received FAS prescriptions and Healthy Start multivitamins (43.4% vs 23.1% and 3.1% vs 1.3%, respectively; p<0.001).

## DISCUSSION

In a study population where 41% of pregnant women had initiated FAS preconception, multiparous women were less likely than primiparous women to do so. Unplanned pregnancies greatly reduced preconceptional FAS initiation, but even among planned pregnancies, two-fifths did not start FAS preconception. Women with infertility or previous pregnancy losses were more likely to start FAS preconception. Nearly half of the women started FAS postconception, mainly after pregnancy recognition at median 5 weeks. For primiparous women with unplanned pregnancies, the first healthcare contact at about 7 weeks gestation further increased FAS initiation. Most women paid for FAS themselves; few obtained it through GP prescriptions, which are free in NI. About a third knew about and ate FA-fortified foods preconception; very few started after conception. Women with poor diets low in folate were less likely to start FAS, despite needing it most.

During preconception, antenatal and interconception care, women are advised on healthy behaviours, including FAS initiation preconception.^24,25^ The higher preconceptional uptake among primiparous women, as well as the increased uptake after pregnancy recognition in our study, suggests that women are not waiting for the antenatal booking appointment to learn about or initiate FAS. These women likely learned about FAS from other sources, including the internet, one of the most common sources of FAS information.^26^

Multiparous women in our study were less likely to initiate FAS preconception, despite their previous pregnancy experience. Previous studies have cited childcare responsibilities^27,28^ or the belief that folic acid was unnecessary given previous healthy pregnancy outcomes,^13,29^ as barriers to early FAS initiation among multiparous women. Our findings suggest otherwise, as multiparous women eventually initiated FAS postconception. We found that women who did not initiate FAS preconception were very likely to do so after pregnancy recognition. FAS initiation following pregnancy recognition indicates some awareness of the importance of folic acid supplementation. As such, our findings suggest the challenge is motivation, not necessarily knowledge. However, it is also possible that women were not sufficiently aware of the importance of the timing of FAS initiation. Moreover, we found that where women reported receiving treatment for infertility, or two or more previous pregnancy losses, they were more likely to initiate FAS preconception, also indicative of motivation. The Capability-Opportunity-Motivation Behaviour framework emphasises that changing behaviour requires improving capability (physical and psychological), opportunity (physical and social), and motivation (automatic and reflective).^30^

Previous studies have largely focused on women’s capability, that is, knowledge and awareness, for FAS uptake.^31^ Our study highlights that motivation needs further exploration when designing public health interventions to increase preconceptional FAS uptake. For example, discussions about maintaining healthy behaviours in future pregnancies and the importance of timing could start in preconception care,^32,33^ and be reinforced in antenatal and interconception care. Such discussions should draw attention to the tendency for multiparous women not to think about FAS until they become pregnant. Opportunity may also be created by widening access to free FAS, either via GP prescriptions (which are free in NI), pharmacies, or via improvements to the Healthy Start scheme (a means-tested scheme in the UK). The Healthy Start scheme has had low uptake among eligible women,^34^ and its eligibility for free vitamins for the mother begins at 10 weeks’ gestation and continues only while breastfeeding. Improving access to free FAS is important in the context of reduced odds of initiating FAS preconception among women living in more deprived areas.

Among women at a higher risk of NTDs, mainly women with obesity in our study population, FAS uptake preconception was not significantly higher than that of low-risk women, despite their greater clinical need. This suggests that more efforts are needed to target high-risk women in NI. We also found that a majority of these high-risk women took the normal (400mcg) dose, with only a few taking the recommended 5mg dose. There is, however, an inconsistency between guidelines which recommend a higher FAS dose for women with obesity,^21,35^ and guidelines which do not^36^ within NI and the UK. This could in part explain the underutilisation of 5mg among high-risk women in this study.

The World Health Organisation advises that populations achieve a minimum red blood cell (RBC) folate level of 906 nanomoles per litre to ensure protection against NTDs.^37^ In the UK, the prevalence of RBC folate insufficiency increased from 69% in 2008 to 89% in 2019 among women of childbearing age.^38^ In our study, very few women intentionally ate FA-fortified foods, although the majority ate at least one fortified brand when provided with a list of brands. This indicates limited awareness of the importance of FA-fortified foods in achieving adequate folate levels to prevent NTDs. This insight matters because fewer than half of the study population took FAS preconception. These findings further reinforce the need for mandatory food fortification. With mandatory fortification starting in December 2026, we expect this to increase RBC folate status in the UK population over time.^39^ Nevertheless, the recommendation for periconceptional FAS will be continued to optimally prevent NTDs.^40^

This study is among the few to focus on parity as a key determinant of FAS initiation preconception, and it is the only one to thoroughly investigate the timing of FAS initiation by parity, pregnancy planning status, timing of pregnancy recognition, and pregnancy-related healthcare. The NI Baby Hearts study captured rich data from a nationally representative sample, making these findings generalisable to the pregnant NI population.^19^ The NI Baby Hearts study also concentrated on periconceptional exposures which were particularly relevant to this study. While the results are robust within the NI context and many findings are consistent with the evidence from other parts of the world, some aspects may be more specific to NI, e.g. the availability of voluntarily fortified foods during the study period and provision of free prescriptions.

While the data-rich, population-based design and the dataset’s representativeness are significant strengths, its origin in a case-control study design is a limitation given our study objective. A longitudinal study in which women are followed from the first to subsequent pregnancies would be most appropriate for achieving our study aims. Instead, our analyses relied on primiparous women’s reported behaviours as a proxy for those of multiparous women during their first pregnancy. Further, this study did not assess the folate content in voluntarily fortified foods consumed by the women.

## CONCLUSION

More women initiated FAS postconception than preconception, and did so particularly after pregnancy recognition at 5 weeks gestation, which is too late for NTD prevention. Multiparous women, despite their previous pregnancy experience, were less likely to initiate FAS preconception than primiparous women. Even among women who planned their pregnancy, two-fifths did not initiate FAS preconception. Initiating FAS after pregnancy recognition suggests some knowledge of its importance; however, this knowledge was insufficient to ensure timely supplementation. Interventions should encourage supplementation within a COM-B framework and pay particular attention to multiparous women. Women living in more deprived areas, as well as those on low-folate diets, were less likely to initiate FAS preconception, even though they needed it most. Such disparities create inequalities in folic acid uptake. The UK Government’s new mandatory FA fortification of non-wholemeal flour could help address these inequalities.

## ACKNOWLEDGEMENTS

This study was funded by Ulster University and the Department for the Economy, Northern Ireland, as part of a PhD studentship. We thank our PPI contributors, Eadaoin O’Kane, Rachel Campbell, and Nicole Winters, whose contributions helped shape this study and interpret its findings.

The NI Baby Hearts study was funded by Northern Ireland Chest Heart and Stroke, with additional support from Children’s Heartbeat Trust. We want to thank the research team, the contributors, and the women who participated in the NI Baby Hearts study. We also wish to acknowledge Nichola McCullough, who structured the NI Baby Hearts dataset and authored publications on the data.

## DATA AVAILABILITY STATEMENT

Due to consent restrictions, the data cannot be publicly shared. Researchers may request access from the corresponding author.

## KEY POINTS

- Previous studies often pointed to knowledge as the reason why women are not initiating folic acid preconception, but our findings suggest motivation as the core challenge.
- Healthcare professionals should embed discussions on pregnancy intention and folic acid use with women, particularly multiparous women, before, during, and after pregnancy.
- Public health interventions should integrate components of the COM-B framework to ensure a holistic approach to encouraging folic acid supplementation.
- Our results on voluntarily folic acid fortified foods reinforce the need for the UK Government’s planned mandatory fortification policy starting December 2026.

