## Supplementary materials for "Timing matters: when do primiparous and multiparous pregnant women initiate folic acid supplementation, and who initiates it too late?"

**STROBE checklist for cross-sectional studies**

| **Item** | **Recommendation** | **Location** |
| --- | --- | --- |
| 1a | Indicate the study design with a commonly used term in the title or abstract | Abstract; Methods |
| 1b | Provide an informative and balanced summary of what was done and what was found | Abstract |
| 2 | Explain the scientific background and rationale | Introduction |
| 3 | State specific objectives, including prespecified hypotheses | Introduction |
| 4 | Present key elements of study design early in the paper | Methods: Study design and participants |
| 5 | Describe setting, locations and dates | Methods: Study design and participants |
| 6a | Eligibility criteria and participant selection | Methods: Study design and participants |
| 7 | Define outcomes, exposures, confounders and modifiers | Methods sections |
| 8 | Sources of data and measurement methods | Methods sections |
| 9 | Efforts to address bias | Statistical analyses |
| 10 | Explain study size | Study design and participants |
| 11 | Handling of quantitative variables | Covariates; Statistical analyses |
| 12a | Statistical methods and confounding control | Statistical analyses |
| 12b | Subgroup and interaction analyses | Statistical analyses |
| 12c | Missing data addressed | Statistical analyses; Tables 1-2 |
| 12d | Sampling strategy | Not applicable |
| 12e | Sensitivity analyses | Not applicable |
| 13a | Numbers at each stage | Results |
| 13b | Reasons for non-participation | Not available from secondary dataset |
| 13c | Flow diagram | Not applicable. |
| 14a | Participant characteristics | Results; Table 1 |
| 14b | Missing data for variables | Tables 1 and 2 |
| 15 | Outcome events or summary measures | Results |
| 16a | Unadjusted and adjusted estimates | Table 2 |
| 16b | Category boundaries | Tables 1 and 2 |
| 16c | Absolute risk translation | Not applicable |
| 17 | Other analyses | Table 3; Supplementary Tables |
| 18 | Key results | Discussion |
| 19 | Limitations | Discussion |
| 20 | Interpretation | Discussion |
| 21 | Generalisability | Discussion |
| 22 | Funding source and role | Funding section |

**Table S1: Definition of study variables**

| **Term** | **Definition** | **Data source** | **Questionnaire item** |
| --- | --- | --- | --- |
| Ate fortified foods in the 3 months after pregnancy | Was drawn from responses to questions about eating fortified breads, breakfast cereals, cereal bars and spreads during the first three months of pregnancy. Women were then categorised based on the number of fortified foods consumed in the first three months of pregnancy: none, one type, or two or more types. | Questionnaire | Q20. During the FIRST THREE MONTHS of your pregnancy which of the following types of folic acid fortified breads did you eat three times a week or more? (Please select all that apply):   - Irwins High fibre Brown - Irwins Sandwich pan white - Irwins Toasty pan white - Irwins Rolls (white/high fibre brown/ white finger) - None of these breads   Q21. During the FIRST THREE MONTHS of your pregnancy which of the following types of breakfast cereals did you eat three times a week or more? (Please select all that apply):   - Muesli or Granola - Porridge - Puffed Wheat - Organic cereals - Shredded Wheat - Selected Kelloggs cereals (see lists on following pages) - Selected Nestle cereals (see lists on following pages) - Selected Weetabix cereals (see lists on following pages) - Selected Asda cereals (see lists on following pages) - Selected Marks and Spencer’s cereals (see lists on following pages) - Selected Sainsburys cereals (see lists on following pages) - Selected Tesco cereals (see lists on following pages) - Other cereals not listed (Go to Q 22) - I don’t eat cereals (Go to Q 22)   Q22. During the FIRST THREE MONTHS of your pregnancy did you eat any of the selected cereal bars (see lists on following pages) three times a week or more? (Please select yes or no):   - Yes - No   Q23. During the FIRST THREE MONTHS of your pregnancy did you use any of the selected types of spreads (see list on following pages) three times a week or more? (Please select yes or no):   - Yes - No |
| Contact with health services | Refers to the gestational week when women first saw a health professional about their pregnancy | Questionnaire | Q4. How many weeks pregnant were you when you first saw a health professional about your pregnancy? |
| Dietary habit | A latent class modelling was used to categorise pregnant women into three dietary classes based on the probability of consuming the different food types three or more times weekly during preconception:   - Type 1 (varied diet class) with high consumption of food types and the highest intake of fruits and vegetables - Type 2 (moderate diet class) with a moderate intake of food types as well as fruits and vegetables - Type 3 (poor diet class) with low intake of food types as well as low fruits and vegetables | Questionnaire | Q24 – 35:  Which of the following foods were you eating during the THREE MONTHS BEFORE you became pregnant?  (Please select one for each type of food):   - Broccoli, brussels sprouts, spinach, peas, dark leafy vegetables - Raw or lightly cooked vegetables - Brown rice, chickpeas, kidney beans, lentils - Oranges, strawberries, raspberries, pineapple, kiwi, cantaloupe, lemons and limes - Other fresh fruits, e.g. apples, bananas, pears, other melons - Tomatoes - Liver - Other fresh meat e.g. beef, chicken - Processed meat e.g. sausages, bacon - Fish - Milk and dairy - Special low calorie foods for dieting |
| Folic acid supplementation | Self-report of taking folic acid supplements or multivitamins/iron supplements containing folic acid, irrespective of the dosage.  Preconceptional FAS was defined as initiating folic acid use before conception.  Postconceptional FAS was defined as folic acid use initiated from the first week of pregnancy. | Questionnaire | Q16. When did you start to take folic acid? (Please select one):   - Before I became pregnant - When I was pregnant   16b. How many weeks pregnant were you when you started to take folic acid?  …………………… weeks |
| High-risk women | Women with pregestational diabetes or epilepsy, or taking anti-epileptic or anti-diabetic medication. Women who were obese (BMI ≥ 30) were also included in this category as per NI guidelines. | NIMATS |  |
| Mental illness | Self-report of having been diagnosed with and still suffering from bipolar disorder, anxiety/stress, depression, obsessive-compulsive disorder, panic disorder and other mental health problems during the current pregnancy.  This also includes women taking medication for any of these conditions. | Questionnaire | Q44. Have you ever been diagnosed by a doctor with any of the following chronic health conditions? (Please select all that apply)  Q46. Are you still suffering from the condition(s)? (Please select one for each condition)  Q51. During the FIRST THREE MONTHS of your pregnancy, did you take/receive any of the following types of medications, supplements, treatments or interventions? (Please select or enter all that apply). For each medication treatment or intervention, please give us the name(s) of all that you  took.  Also, tell us when you first started taking this type of medication, treatment or intervention, when you stopped, or if you are still taking/receiving it. |
| Parity  *Primiparity*  *Multiparity* | Regardless of the outcome, the number of viable pregnancies per woman (viability referring to pregnancies that are at least 24 gestational weeks 0 days):  *Women carrying their first viable pregnancy and coded in the maternity database as 0 + …, where the first digit (0) connotes primiparity and the second connotes previous pregnancy loss.*  *Women with a previous viable pregnancy and coded in the maternity database as 1 + …, 2 + …, etc.* | NIMATS |  |
| Pregnancy planning | Women who self-reported trying to get pregnant | Questionnaire | Q12. How long were you trying to get pregnant? (Please select one)   - I was not planning to become pregnant - Up to one year - Longer than a year |
| Pregnancy recognition  *Pregnancy suspicion*  *Pregnancy confirmation* | Refers to period spanning from when women suspected and confirmed that they were pregnant.  *This is gestational week when the woman reported first suspecting that they might be pregnant.*  *This is gestational week when the woman reported confirming that she is pregnant.* | Questionnaire | *Q2. How many weeks pregnant were you when you first thought you might be pregnant?*  *Q3. How many weeks pregnant were you when you had your pregnancy confirmed?* |
| Pregnancy-related stress during the three months before pregnancy | Self-report of feeling stressed about any aspect of their pregnancy three months before conception | Questionnaire | Q91. Have you been stressed about any aspect of your pregnancy, or about becoming pregnant?   - No - Yes, during the THREE MONTHS BEFORE I became pregnant - Yes, during the FIRST THREE MONTHS of pregnancy - Yes, more recently |
| Selectively buying fortified foods | Assessed from questionnaire responses to question 19 | Questionnaire | Q19. Do you buy cereals, breads, cereal bars or spreads fortified with folic acid? (Please select one):   - Yes- since **before** I became pregnant - Yes- since I **became** pregnant - No - I don’t know |
| Stressful events during the three months before pregnancy | Self-report on conditions during the three months before conception. | Questionnaire | Options for Q81-90:   - No - Yes, during the THREE MONTHS BEFORE I became pregnant - Yes, during the FIRST THREE MONTHS of pregnancy - Yes, more recently   Q81. Have you experienced the death(s) of an immediate member of the family, other family member or a close friend?  Q82. Have you or a close family member or friend had a serious illness or injury?  Q83. Have you moved house?  Q84. Have you or your partner had serious trouble at work or become unemployed?  Q85. Have you had major relationship difficulties with your partner or husband, or become separated or divorced?  Q86. Has someone close to you experienced substance abuse or alcohol problems?  Q87. Have you experienced social or ethnic discrimination?  Q88. Have you or your partner had serious legal or financial problems?  Q89. Have you or anyone close to you been a victim of violence or crime, including domestic violence?  Q90. Have you or anyone close to you been arrested or held in prison? |

More information is available on [The Baby Hearts Study (Northern Ireland) Database - Ulster University](https://pure.ulster.ac.uk/en/datasets/the-baby-hearts-study-northern-ireland-database/).

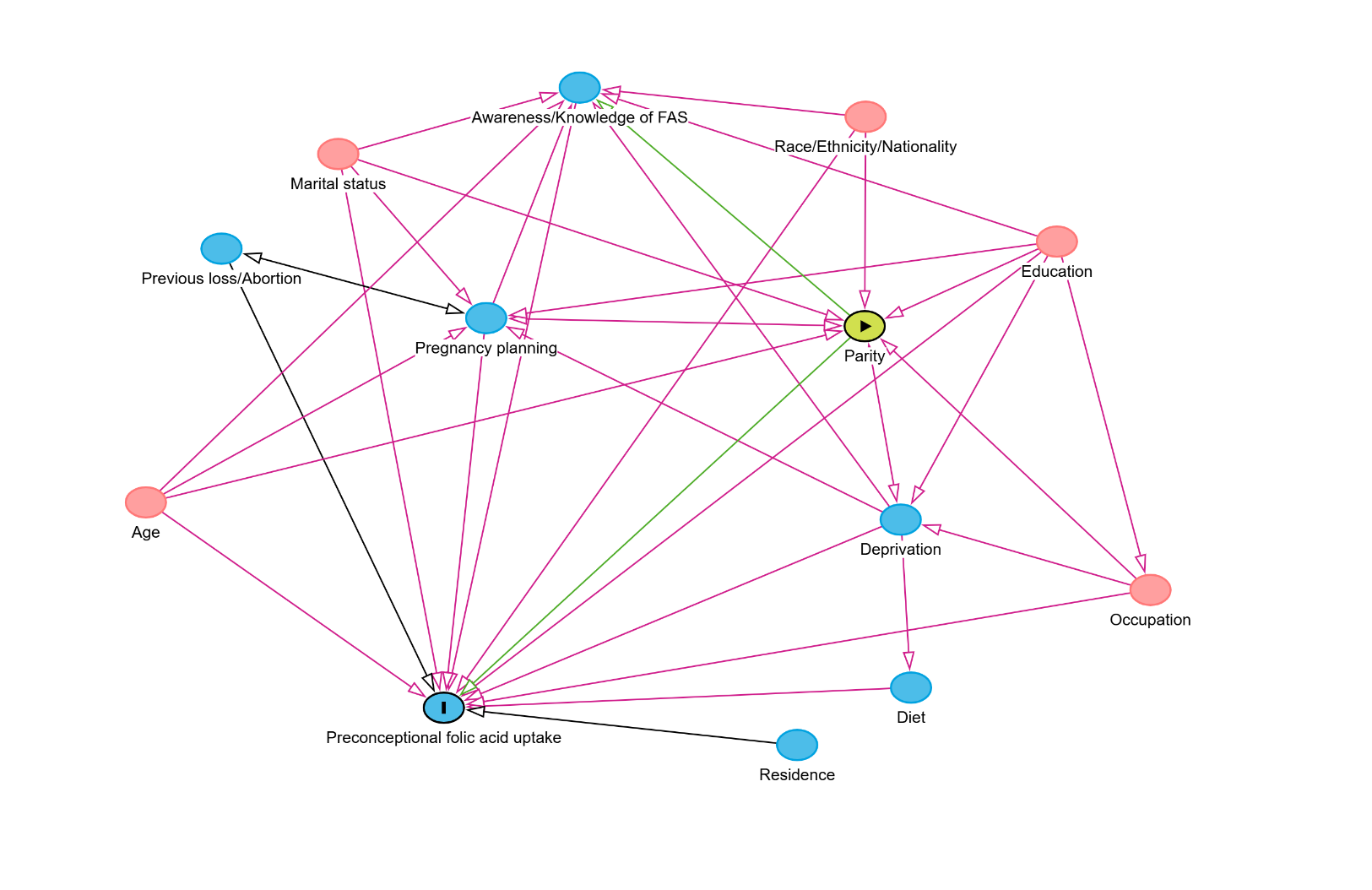
**Figure S1: Evidence from literature on the relationship between DAG variables**

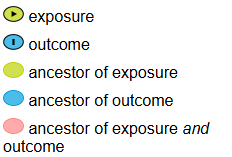
**Legend**

**Table S2: Evidence from literature on the relationship between DAG variables**

| **Outcomes** | **Associated determinants** | **References** |
| --- | --- | --- |
| Folic acid uptake | Age, parity and pregnancy planning | ^1–8^ |
|  | Education, Marital status, and Deprivation | ^1,9–13^ |
|  | Race/ethnicity or Nationality | ^1,4,12,14^ |
|  | Awareness or knowledge of folic acid | ^1,15^ |
|  | Occupation | ^3,11,16–18^ |
|  | Residence (urban vs rural) | ^11,16,19^ |
|  | Contact with health services (Antenatal care) | ^16,17,19^ |
|  | Previous pregnancy loss or abortion | ^19^ |
|  | Diet | ^11^ |
| Awareness or knowledge of folic acid | Pregnancy planning, Timing when pregnancy was recognised, Education, Marital status, Parity, and Age | ^13^ |
|  | Race/ethnicity, Education, Marital status, Deprivation | ^15^ |
|  | Age, Education, and Marital Status | ^20^ |
| Parity | Age, Marital status, and Timing of health service contact | ^21^ |
|  | Education, Occupation, Age, Deprivation, and Marital status | ^22^ |
|  | Race/Ethnicity, and Education | ^23^ |
|  | Pregnancy planning | ^24–27^ |
| Pregnancy planning | Age, Parity, Deprivation, and Previous abortion | ^28^ |
|  | Age, Parity, Education and Deprivation | ^27^ |
|  | Marital status, Parity, and Folic acid uptake | ^25^ |

**Table S3: Folic acid and dietary intake by parity and pregnancy planning**

|  | **Primiparous women**  **(n = 393)** | | | **Multiparous women**  **(n = 573)** | | | **All women**  **(n = 966)** | |
| --- | --- | --- | --- | --- | --- | --- | --- | --- |
|  | **Unplanned pregnancy**  **(n = 136)** | **Planned pregnancy**  **(n = 257)** | **p-value** | **Unplanned pregnancy**  **(n = 184)** | **Planned pregnancy**  **(n = 389)** | **p-value** | **Unplanned pregnancy**  **(n = 320)** | **Planned pregnancy**  **(n = 646)** |
| Folic acid intake  Did not take folic acid  Initiated preconception  Initiated postconception | 23 (16.9%)  8 (5.9%)  105 (77.2%) | 23 (8.9%)  168 (65.4%)  66 (25.7%) | **<0.001** | 40 (21.7%)  13 (7.1%)  131 (71.2%) | 30 (7.7%)  205 (52.7%)  154 (39.6%) | **<0.001** | 63 (19.7%)  21 (6.6%)  236 (73.7%) | 53 (8.2%)  373 (57.7%)  220 (34.1%) |
| Folic acid compliance  Everyday  3 to 4 times a week  1 to 2 times a week  1 to 2 times a month  *Missing* | 106 (77.9%)  4 (3.0%)  3 (2.2%)  -  *23 (16.9%)* | 227 (88.3%)  6 (2.3%)  2 (0.8%)  1 (0.4%)  *21 (8.2%)* | 0.49 | 131 (71.2%)  7 (3.8%)  6 (3.3%)  1 (0.5%)  *39 (21.2%)* | 330 (84.8%)  23 (5.9%)  7 (1.8%)  1 (0.3%)  *28 (7.2%)* | 0.34 | 237 (74.1%)  11 (3.4%)  9 (2.8%)  1 (0.3%)  *62 (19.4%)* | 557 (86.2%)  29 (4.5%)  9 (1.4%)  2 (0.3%)  *49 (7.6%)* |
| Women at a higher risk of NTDs˟  Postconception or No intake  Preconception (any dose) | 24 (88.9%)  3 (11.1%) | 15 (31.9%)  32 (68.1%) | **<0.001** | 49 (35.9%)  5 (18.1%) | 44 (51.2%)  42 (48.8%) | **<0.001** | 73 (90.1%)  8 (8.9%) | 59 (44.4%)  74 (55.6%) |
| Selectively bought fortified foods  Yes, since before I became pregnant  Yes, since I became pregnant  No  I don’t know | 35 (25.7%)  11 (8.1%)  57 (41.9%)  33 (24.3%) | 85 (33.1%)  13 (5.1%)  74 (28.7%)  85 (33.1%) | **0.02** | 64 (34.8%)  16 (8.7%)  51 (27.7%)  53 (28.8%) | 150 (38.6%)  16 (4.1%)  132 (33.9%)  91 (23.4%) | **0.04** | 99 (30.9%)  27 (8.4%)  108 (33.8%)  86 (26.9%) | 235 (36.4%)  29 (4.5%)  206 (31.9%)  176 (27.2%) |
| Fortified foods (first trimester)  None  One food type  Two or more food types | 26 (19.1%)  53 (39.0%)  57 (41.9%) | 57 (22.2%)  90 (35.0%)  110 (42.8%) | 0.67 | 33 (17.9%)  83 (45.1%)  68 (37.0%) | 77 (19.8%)  149 (38.3%)  163 (41.9%) | 0.29 | 59 (18.4%)  136 (42.5%)  125 (39.1%) | 134 (20.7%)  239 (37.0%)  273 (42.3%) |
| Dietary habits preconception  Type 1 – Moderate diet  Type 2 – Varied diet  Type 3 – Poor diet  *Missing* | 43 (31.6%)  40 (29.4%)  51 (37.5%)  *2 (1.5%)* | 109 (41.4%)  93 (36.2%)  55 (21.4%)  - | **<0.01** | 76 (41.3%)  39 (21.2%)  69 (37.5%)  - | 150 (38.6%)  154 (39.5%)  85 (21.9%)  - | **<0.001** | 79 (24.7%)  119 (37.2%)  120 (37.5%)  *2 (0.6%)* | 247 (38.2%)  259 (40.1%)  140 (21.7%)  -- |
| ˟ Women with a higher risk do not sum up to the column totals because the analysis only included women at a higher risk of having children with NTDs. | | | | | | | | |

**Table S4: Stratified analysis of the determinants of preconceptional folic acid intake by parity and interaction between parity and determinants**

|  | **Primiparous women**  **(n = 393)** | | | **Multiparous women**  **(n = 573)** | | | **Interaction p-value** |
| --- | --- | --- | --- | --- | --- | --- | --- |
|  | **PFAS**  **(n = 176)** | **NFAS**  **(n = 217)** | **Odds Ratio**  **(95% CI)** | **PFAS**  **(n = 218)** | **NFAS**  **(n = 355)** | **Odds Ratio**  **(95% CI)** |  |
| Maternal occupation  Employed  Unemployed  Homemakers  Students | 172 (48.9%)  2 (9.1%)  2 (50.0%)  -- | 180 (51.1%)  20 (90.9%)  2 (50.0%)  15 (100.0%) | Ref  **0.11 (0.02 – 0.46)**  1.05 (0.15 – 7.51)  -- | 185 (44.4%)  4 (8.7%)  29 (27.9%)  -- | 232 (55.6%)  42 (91.3%)  75 (72.1%)  5 (100.0%) | Ref  **0.12 (0.04 – 0.34)**  **0.49 (0.30 – 0.78)**  -- | 0.75 |
| Area-based deprivation  1 (most deprived)  2  3  4  5 (least deprived)  *Missing* | 21 (27.3%)  33 (42.3%)  49 (58.3%)  38 (47.5%)  32 (50.0%)  *7 (70.0%)* | 56 (72.7%)  45 (57.7%)  35 (41.7%)  42 (52.5%)  32 (50.0%)  *3 (30.0%)* | **1.24 (1.07 – 1.44)** | 33 (27.7%)  42 (34.4%)  53 (41.4%)  37 (33.3%)  48 (57.8%)  *5 (50.0%)* | 86 (72.3%)  80 (65.6%)  75 (58.6%)  74 (66.7%)  35 (42.2%)  *5 (50.0%)* | **1.27 (1.11 – 1.44)** | 0.83 |
| Infertility treatment  None  Investigations only  Treated for infertility  *Missing* | 118 (37.3%)  11 (61.1%)  33 (94.3%)  *10 (41.7%)* | 198 (62.7%)  7 (38.9%)  2 (5.7%)  *14 (58.3%)* | Ref  2.64 (0.99 – 6.99)  **27.69 (6.53 – 117.49)** | 198 (37.6%)  4 (36.4%)  5 (41.7%)  *11 (45.8%)* | 328 (62.4%)  7 (63.6%)  7 (58.3%)  *13 (54.2%)* | Ref  0.95 (0.27 – 3.28)  1.18 (0.37 – 3.78) | **0.002** |
| Previous pregnancy loss  None  1  2 or more  *Missing* | 128 (41.3%)  30 (50.8%)  10 (83.3%)  *4 (33.3%)* | 182 (58.7%)  29 (49.2%)  2 (16.7%)  *8 (66.7%)* | Ref  1.47 (0.84 – 2.57)  **7.11 (1.53 – 32.99)** | 107 (33.8%)  68 (41.2%)  35 (46.7%)  *8 (50.0%)* | 210 (66.2%)  97 (58.8%)  40 (53.3%)  *8 (50.0%)* | Ref  1.38 (0.93 – 2.03)  **1.72 (1.03 – 2.86)** | 0.23 |
| Pregnancy planning  Unplanned pregnancy  Planned pregnancy | 8 (5.9%)  168 (65.4%) | 128 (94.1%)  89 (34.6%) | **0.03 (0.02 – 0.07)**  Ref | 13 (7.1%)  205 (52.7%) | 171 (92.9%)  184 (47.3%) | **0.07 (0.04 – 0.12)**  Ref | 0.14 |
| Diet before pregnancy  Type 1 – Moderate diet  Type 2 – Healthy diet  Type 3 – Poor diet  *Missing* | 77 (50.7%)  71 (53.4%)  28 (26.4%)  *2 (100.0%)* | 75 (49.3%)  62 (46.6%)  78 (73.6%)  *--* | 0.89 (0.56 – 1.43)  Ref  **0.31 (0.18 – 0.54)** | 81 (35.8%)  102 (52.8%)  35 (22.7%)  -- | 145 (64.2%)  91 (47.2%)  119 (77.3%)  -- | **0.49 (0.34 – 0.74)**  Ref  **0.26 (0.16 – 0.42)** | 0.16 |
| *PFAS = Preconceptional initiation; NFAS = Postconceptional initiation or No intake* | | | | | | | |
